# Potential targets for the treatment of MI: GRP75-mediated Ca^2+^ transfer in MAM

**DOI:** 10.1101/2023.10.17.23297179

**Authors:** Chenyan Zhang, Bowen Liu, Jiaxing Sheng, Jia Wang, Weijie Zhu, Chen Xie, Xuan Zhou, Yuxin Zhang, Qinghai Meng, Yu Li

## Abstract

**Background:** After myocardial infarction (MI), there is a notable disruption in cellular calcium ion homeostasis and mitochondrial function. These alterations are believed to be linked to endoplasmic reticulum (ER) stress, though the specific mechanisms are not fully understood. This research endeavors to elucidate the involvement of glucose regulated protein 75 (GRP75) in post-MI calcium ion homeostasis and mitochondrial function.

**Results:** Excessive oxidative stress was activated in humans’ post-myocardial infarction, with most differentially expressed genes being enriched in metabolic pathways, especially the calcium signaling pathway. In MI rats, symptoms of myocardial injury were accompanied by an increase in the activation of PERK, ATF6, and IRE1, as well as elevated Binding immunoglobulin protein (Bip) expression. Moreover, in oxygen-glucose deprivation (OGD)-induced cardiomyocytes, it was confirmed that inhibiting PERK exacerbated intracellular Ca^2+^ disruption and cell apoptosis. More importantly, in cardiomyocytes undergoing Tunicamycin-induced ER stress, Ca^2+^ accumulated in both the ER and mitochondria. Concurrently, the co-localization of GRP75 with IP3R and VDAC1 increased under ER stress in cardiomyocytes. In OGD-induced cardiomyocytes, knockdown of GRP75 not only reduced the Ca^2+^ levels in both the ER and mitochondria and improved the ultrastructure of cardiomyocytes, but it also increased the number of contact points between ER and mitochondria, reducing MAM formation, and decreased cell apoptosis. Significantly, knockdown of GRP75 did not affect the protein expression of PERK and hypoxia-inducible factor 1α (HIF-1α). Transcriptome analysis of cardiomyocytes revealed that knockdown of GRP75 mainly influenced the molecular functions of sialyltransferase and IP3R, as well as the biosynthesis of glycosphingolipids and lactate metabolism. In OGD-induced cardiomyocytes, the knockdown of GRP75 lowered the protein expression levels of glucose transporter-1 (Glut1), pyruvate kinase M2 (PKM2), and lactate dehydrogenase A (LDHA), and decreased the metabolic products of glycolysis.

**Conclusion:** The complex interaction between the ER and mitochondria, driven by the GRP75 and its associated IP3R1-GRP75-VDAC1 complex, is crucial for calcium homeostasis and cardiomyocyte’s adaptive response to ER stress. Modulating GRP75 could offer a strategy to regulate calcium dynamics, diminish glycolysis, and thereby mitigate cardiomyocyte apoptosis.

## Introduction

Cardiovascular disease imposes a huge burden on global health systems^1^. Various cardiovascular diseases have high mortality and rehospitalization rates. The endoplasmic reticulum (ER) is a key organelle that regulates intracellular material transport. It plays a crucial role in the secretion and folding of membrane proteins^2^, posttranslational modification, and regulation of calcium homeostasis. The ER quality control system (ERQC) can facilitate the correct folding of proteins as well as selectively attack incorrectly folded polypeptides^3^. Such a working mission allows a tight balance between intracellular Ca^2+^, molecular chaperones, and oxidative stress in the lumen of the ER^4^. When energy dyshomeostasis, changes in Ca^2+^ levels and oxidative stress occur in cells, unfolded or misfolded proteins accumulate in the ER, which is known as ER stress^5^. Stimuli from the internal and external environments, such as ischemia, hypoxia and oxidative stress, are important factors that trigger ERs in myocardial infarction (MI)^6^. Ca^2+^ may be a key regulator in the association of ERs with MI^7^. Circulatory disturbance of Ca^2+^ leads to the occurrence of MI and the decline of myocardial contractile function.

Calcium ion homeostasis in mitochondria is essential for key enzymes of glucose metabolism and respiratory chain complexes^8^. Excessive uptake of Ca^2+^ by mitochondria can lead to mitochondrial dysfunction and initiate apoptotic pathways^9^. The structure of the physical connection that exists between the ER and mitochondria, called the mitochondria associated endoplasmic reticulum membrane(MAM)^10^, is a dynamic connection made up of subdomains of the ER, the outer mitochondrial membrane, and a series of proteins ^11–13^. ER releases Ca^2+^, mediated by inositol1,4,5-trisphosphate (IP3Rs) or ryanodine receptors (RyRs)^14^. 75% of its released Ca^2+^ are pumped back by the ER sarcoplasmic reticulum calcium pump (SERCA2a), and mitochondria can take up 20% of the calcium ions to maintain intracellular calcium homeostasis through voltage-dependent anion-selective channel (VDAC) which located on the outer mitochondrial membrane (OMM) and mitochondria calcium uniporter (MCU) which located on the inner mitochondrial membrane (IMM). Studies from Szabadkai’s group^15^ have shown that VDAC1 is physically linked to IP3R1 through MAM associated glucose regulated protein 75 (GRP75). GRP75 was first characterized in mammals as a heat shock protein-70 (HSP70) family stress chaperone protein based on its sequence homology. Numerous studies in mammals have shown that GRP75 can be induced by various stressors such as glucose deprivation, oxidative stress, and hypoxia. The latest study^16^ demonstrated that GRP75 directly regulates calcium transfer from ER to mitochondria. The important role of GRP75 in the transmission of Ca^2+^ and mitochondrial dysfunction has been demonstrated in diabetes retinopathy^17^, nephropathy^18^, and neurological diseases^19^, but its mechanism remains to be verified in MI.

The effect of ER stress induced by MI on disease progression remains controversial^20–22^. How mitochondria communicate with the ER affects the myocardium is not yet distinct. We hypothesized that there exists a pathway, IP3R/GRP75/VDAC1, during MI in which over-activation of ER stress leads to abnormal mitochondrial function, which in turn leads to oxygen glucose deprivation (OGD) injury in cardiomyocytes. In this study, we verified that ER stress was activated after MI and we used OGD to simulate the disease environment in cells. The effects of OGD-induced ER stress on mitochondrial function, apoptosis, and myocardial ultrastructure were evaluated. Referring to the central location of GRP75 in MAM outlined earlier, we investigated whether the transfer of Ca^2+^ from ER to mitochondria through the IP3R/GRP75/VDAC1 complex may play a role in mitochondrial calcium overload and subsequent cardiomyocyte death in the OGD model. Our findings provide new ideas and intervention targets for the prevention and treatment of MI.

## Materials and methods

### Major Resources

Major resources were all provided in Supplementary Materials (Table S1 and Table S2).

### GEO database sequencing analysis

Login to the GEO dataset website https://www.ncbi.nlm.nih.gov/gds/. Using the keyword “myocardial infarction” to retrieve the dataset of human samples, after selecting the GSE83500 dataset, 11 samples were selected from the dataset, including 5 samples (GSM2204587, GSM2204591, GSM2204593, GSM2204597, GSM2204618) aortic sample from stable angina (non-MI), and 6 samples (GSM2204584, GSM2204598, GSM2204599, GSM2204607, GSM2204609, GSM2204615) aortic sample from unstable angina (MI). Restricted to Chinese men. Using the official GEP2R analysis tool, define groups (non-MI and MI) according to the sample numbers mentioned above, and perform analysis to obtain differential gene expression profiles, screen differential genes, and perform differential gene KEGG signal enrichment analysis (https://david.ncifcrf.gov/home.jsp). With the R package, we drew a volcano plot and a signal enrichment plot for the screening results of differentially expressed genes.

### Experimental animals

This study was conducted in accordance with internationally accepted animal welfare guidelines and passed the ethical review of Nanjing University of Chinese Medicine (No. 201906A030).20 rats purchased from Hangzhou Medical College [License No. SCXK(Zhe)2019-0002; Zhejiang, China]. The rats were fed full nutritive pellet feed and drank freely. The experiment was carried out after 1 week of adaptive feeding. Adult male Sprague-Dawley rats, 10-weeks-old weighing 300-350 g, were randomly divided into 2 groups, Sham (n=10) and Ischemic (n=10). Animals were kept on a 12/12 h light/dark cycle and received water and food *ad libitum*.

### Modeling and treatment

Rats were anesthetized by intraperitoneal injection of 2ml/kg 2% pentobarbital sodium, according to body weight. After the rats were fixed, electrocardiogram electrodes were connected subcutaneously to the limbs, and endotracheal intubation was connected to the small animal ventilator. Respiratory rate: 90 times /min, tidal volume of 10∼12 mL, respiratory ratio of 1:2. The left anterior chest was sheared and disinfected with iodoprene. A longitudinal incision of about 1 cm was made between the 3rd and 4th ribs on the left side of the sternum to bluntly separate the muscle and pleura. A hemostatic forceps was used to open the third intercostal space and a dilator was placed to secure the incision to expose the surgical field. The 6/0 suture needle was inserted into the lower margin of the left atrial appendage at a depth of about 1.5 mm and a needle distance of about 3-4 mm. The anterior descending branch of the left coronary artery (LAD) was ligated. Myocardial infarction was confirmed by the up-shift of J-point in the electrocardiogram of rats after ligation, and then the incision was sutured layer by layer. For the sham operation group, thread was only used to pass under the LAD after thoracotomy without ligature. For the MI group, a slight whiteness was observed in the precardiac area after the suture was tightened. Wait for the rats to wake up on the blanket before returning to their cages.

### Hemodynamic detection

The anesthetized rats were fixed on the operating table and electrocardiogram and heart rate were recorded with PowerLab physiological recorder (ADI, Australia). A longitudinal skin incision was made along the anterior median line of the neck to bluntly separate the muscle from the fascia and expose the left common carotid artery. The distal end of the heart was tied with cotton thread and the catheter was placed along the common carotid artery and ascending aorta into the left ventricle. Left ventricular systolic blood pressure (LVSP), left ventricular end-diastolic blood pressure (LVEDP), and the maximum rate of rise and fall in the left ventricle (±dp/dt_max_) were recorded.

### Immunoblotting analysis

The concentration of proteins was detected by BCA assay kit (Vazyme, China) according to the instructions. Then 30 μg of protein was separated by 10% SDS-PAGE and transferred to a PVDF membrane (Millipore, U.S.). After blocking the non-specific sites, the primary antibody was incubated with the membrane at 4°C overnight. The membrane was then incubated with the secondary antibody for 120 min. Specific information is shown in Supplementary Table S1. The membranes were then incubated with ECL (Vazyme, China) for luminescence generation. The image and gray level were analyzed with ImageLab software (Bio-Rad, U.S.).

### Immunofluorescence staining

Cells were seeded in 24-well plates and fixed using 4% paraformaldehyde at the end of treatment. After sealing the non-specific binding sites, the proteins on the cells were labelled using primary antibodies from different sources and incubated overnight at 4°C. Secondary antibodies of different fluorescent colors were used to bind to the corresponding primary antibodies. The position of the cell nucleus was labelled with Dapi (Beytime, China). Confocal microscope shots (Leica, Germany).

### Real-time polymerase chain reaction

The mRNA expression of key enzymes of glycolysis was detected by quantitative Real-time polymerase chain reaction (RT-PCR). Total RNA from H9c2 cells was isolated using TRIzol reagent (Invitrogen, U.S.). cDNA was reverse transcribed from RNA using 1st Strand cDNA Synthesis Kit (Vazyme, China), the primer sequences are shown in **Supplymentary Table S2**. Real-time PCR amplification was performed using Taq Pro Universal SYBR qPCR Master Mix (Vazyme Company, China).

### siRNA silencing

Three pairs of GRP75 small interfering RNA (si-RNA) sequences and one pair of non-specific RNA sequences were designed and synthesized by biotechnology company (Kaiji, China). The siRNA was mixed with the transfection reagent and added to the 6-well plate at a concentration of 50 nM (5×10∧5cells/well). Cells were treated at 37° with 5 % CO_2_ for 24 h.

### Transcriptome sequencing

Total RNA was extracted from rat cardiomyocytes H9c2, and the quality of the total RNA was tested. After purification of the mRNA, fragmented mRNA was obtained. After cDNA synthesis, a library was established and quality tested. The second-generation Illumina sequencing platform was used for sequencing.

### LC-MS/MS Detection

The peptides in the samples were separated using a nano-UPLC liquid chromatography system and then coupled to a mass spectrometer for data acquisition. A Reprosil-Pur 120 C18-AQ column (15 cm × 100 μm, 1.9 μm) was used for chromatographic separation. The mass spectrometry analysis was performed in data-dependent acquisition mode and positive ion detection mode, with a primary scanning range of m/z 350-1600 and a resolution of 120 000 @m/z 200; the AGC was 3E6, and the maximum ion implantation time was 30 ms. The secondary scanning was fixed at a minimum m/z of 110 and a resolution of 45 000. Based on the peak width of the chromatographic peaks, the dynamic exclusion time was set to 45 s. The dynamic exclusion time of the chromatographic peaks was set to 45 s.

### Statistical analysis

All data were expressed as the Mean ± Standard error of the mean (SEM). Each figure legends contains detailed information on sample size and data collection. Data were analyzed using either the two-tailed unpaired Student t-test or the non-parametric Mann-Whitney U test to compare means between the two experimental groups; analysis of variance (ANOVA) followed by the Bonferroni post-hoc test was used to compare means among multiple groups where appropriate; Statistical analyses were performed using GraphPad Prism (Version 9.0; GraphPad Software, Inc., USA). *p* < 0.05 was considered statistically significant.

## Results

### Endoplasmic reticulum stress is activated after myocardial infarction

Extracting the Chinese male transcriptome sequencing data from the GEO database (GSE83500), grouping them into non-MI and MI, and standardizing the data (**Figure 1A**). The analysis of the sample background information revealed that there was no significant difference in the age of the two group (**Figure 1B**). The volcano map indicates the genes that were up-regulated and down-regulated in male aortic smooth muscle cells after MI (**Figure 1C**). Kyoto Encyclopedia of Genes and Genomes (KEGG) signal enrichment analysis of differentially expressed genes between the two groups revealed that most differentially expressed genes in MI were enriched in metabolic pathways (**Figure 1D**). The change in metabolic pathways ranks first among all significant signal pathways, with the calcium signaling pathway playing an important role in MI (**Figure 1E-F**).

**Figure 1.**
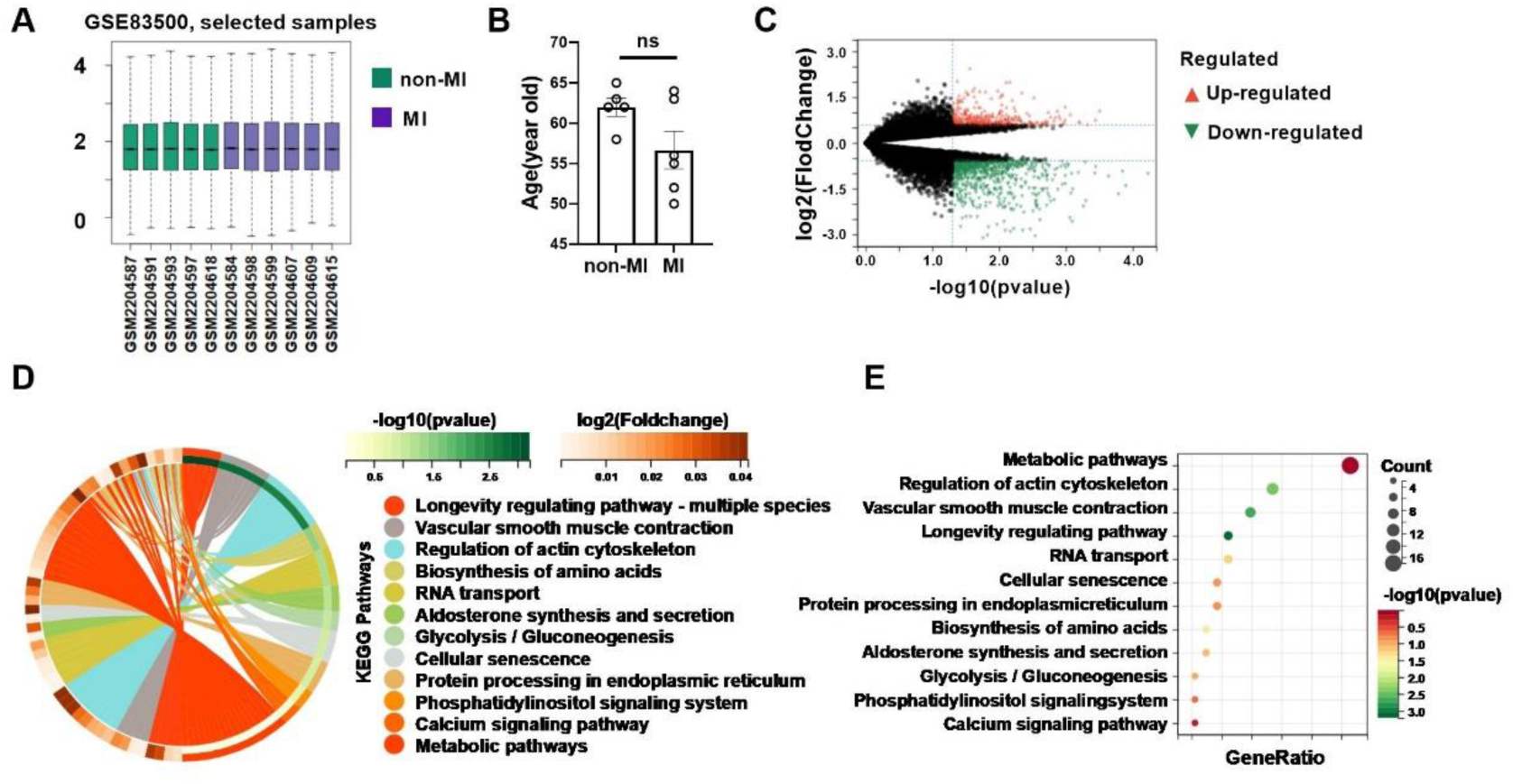
Transcriptome analysis of aortic vascular smooth muscle cells in patients before and after Myocardial Infarction. Data were extracted from the GSE83500 dataset in GEO for further analysis. The sample type was aortic vascular smooth muscle cells from patients before and after MI, and the data type was transcriptome sequencing data. **(A)** Information grouping and data standardization. **(B)** Age data were extracted from the background data to analyze the difference between the two groups. **(C)** The differentially expressed genes between the two groups were analyzed and a volcano plot was drawn. KEGG signal enrichment was performed on the differentially expressed genes between the two groups, and a circle plot **(D)** and bubble plot **(E)** were drawn. Values are the mean ± SEM, n = 5 in non-MI group, n = 6 in MI group, two-tailed Student’s t-test.

Next, we reproduced the MI rat model by ligating the left anterior descending artery, and observed the electrocardiogram (ECG) at the time of surgery to evaluate whether the modeling was successful or not. In the ischemic group, J-point elevation was accompanied by the appearance of pathological Q waves (**Figure 2A**), representing the success of model replication. 2,3,5-triphenyl tetrazolium chloride (TTC) staining was used to verify MI after operation (**Figure 2B**), and the degree of myocardial fibrosis was observed in the long term (**Figure 2C**). Hemodynamics of left ventricle and femoral artery in rats were monitored (**Figure 2D-H**). The contents of CK-MB and BNP in serum were detected (**Fig. 2I-J**). In MI mice, the activated three signaling pathways: protein kinase R-like endoplasmic reticulum kinase (PERK), activating transcription factor 6 (ATF6), and inositol requiring enzyme 1 (IRE1) were compared by the up-regulation of Binding immunoglobulin protein (Bip) protein expression (**Figure 2K**). Notably, there was a more pronounced activation of the PERK pathway (**Figure 2L-O**).

**Figure 2.**
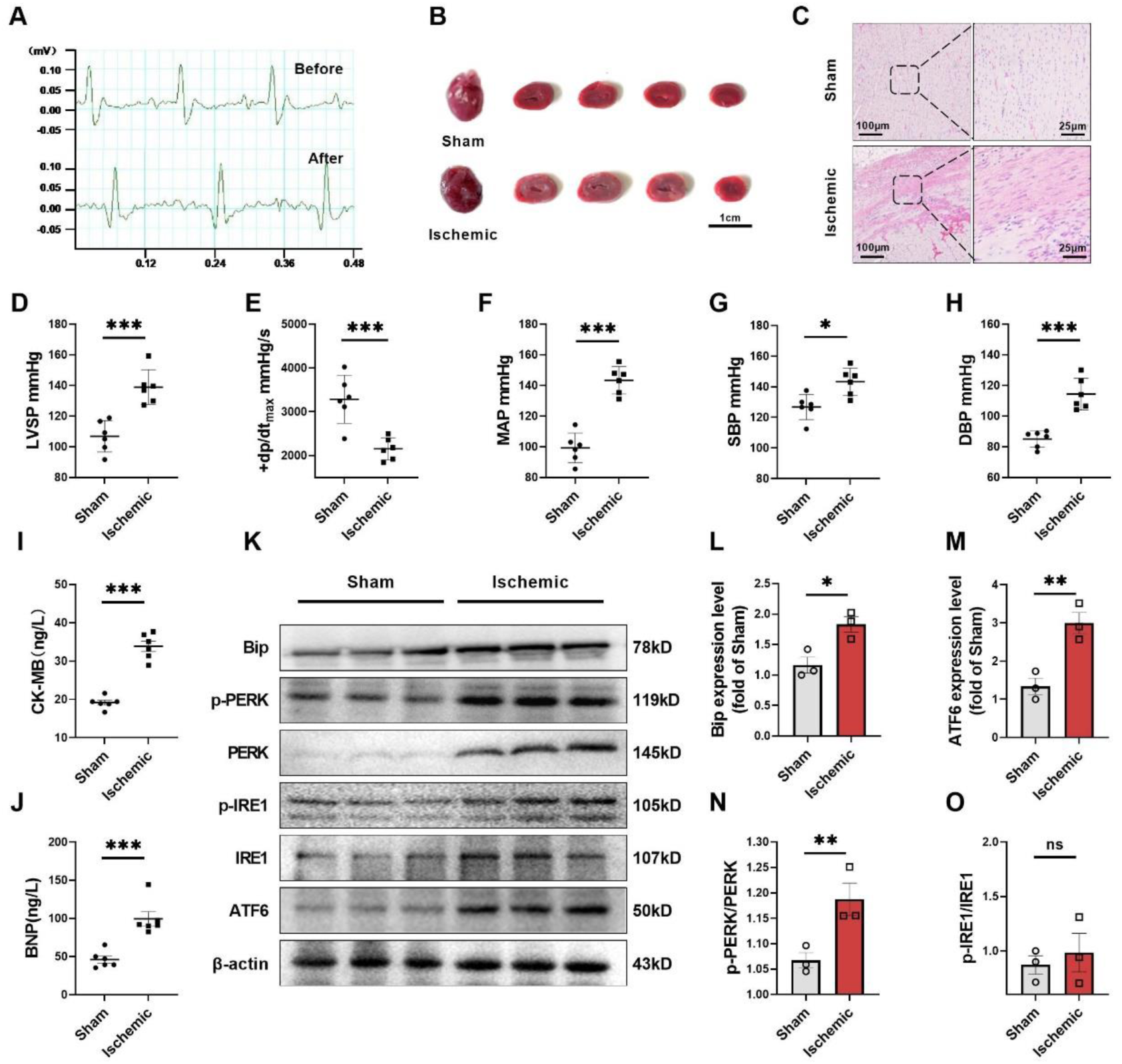
Endoplasmic reticulum stress is activated after myocardial infarction The electrocardiogram changes were used as the intraoperative evaluation criteria. (A). Myocardial ischemic area was assessed by 2%TTC staining after surgery (B). The degree of fibrosis was observed at long term using Sirius red staining (C). Left ventricular systolic pressure (D), maximum rise rate of left indoor pressure (E), mean femoral arterial pressure (F), systolic blood pressure (G), and diastolic blood pressure (H) were recorded. Serum changes of CK-MB (I) and BNP (J) were measured 24 hours after surgery. Values are the mean ± SEM. n = 6 per group. \**P* < 0.05, \*\*\**P* < 0.001, two-tailed Student’s t-test. Immunoblotting detected ER stress protein Bip, p-PERK, PERK, p-IRE1, IRE1, and MCU levels in rat myocardium (K-O). Values are the mean ± SEM. n = 3 per group. \**P* < 0.05, \*\**P* < 0.01, two-tailed Student’s t-test.

### Protective role of PERK activation against OGD-induced cardiomyocyte apoptosis

To address the activation of ER stress after MI, we used three different inhibitors to treat H9c2. GSK2656157 (PERK inhibitor), STF-083010 (IRE1 inhibitor), and AEBSF (ATF6 inhibitor). We found that when PERK was inhibited, greater damage occurred in the face of OGD injury (**Figure 3A**). Unlike the damage caused by OGD, flow cytometry detection revealed that inhibition of the PERK pathway leads to more significant cell apoptosis (**Figure 3B-C**). Both mitochondrial membrane potential and ATP content showed significant decreases after OGD, and this situation was aggravated by PERK inhibitors (**Fig. 3D-F**). When intracellular Ca^2+^ concentration was detected, it was found that a significant elevation of intracellular Ca^2+^ concentration occurred in the PERK inhibitor group compared to the control group, and OGD exacerbated such calcium exocytosis (**Fig. 3G**). A significant amount of intracellular calcium was exported to the extracellular space in the model group (**Figure 3H**). Immunoblotting detects more significant damage in the apoptosis of cells (**Figure 3I-K**). In summary, PERK activation can prevent cell apoptosis caused by OGD. The treatment of PERK inhibitors exacerbated the apoptosis induced by OGD.

**Figure 3.**
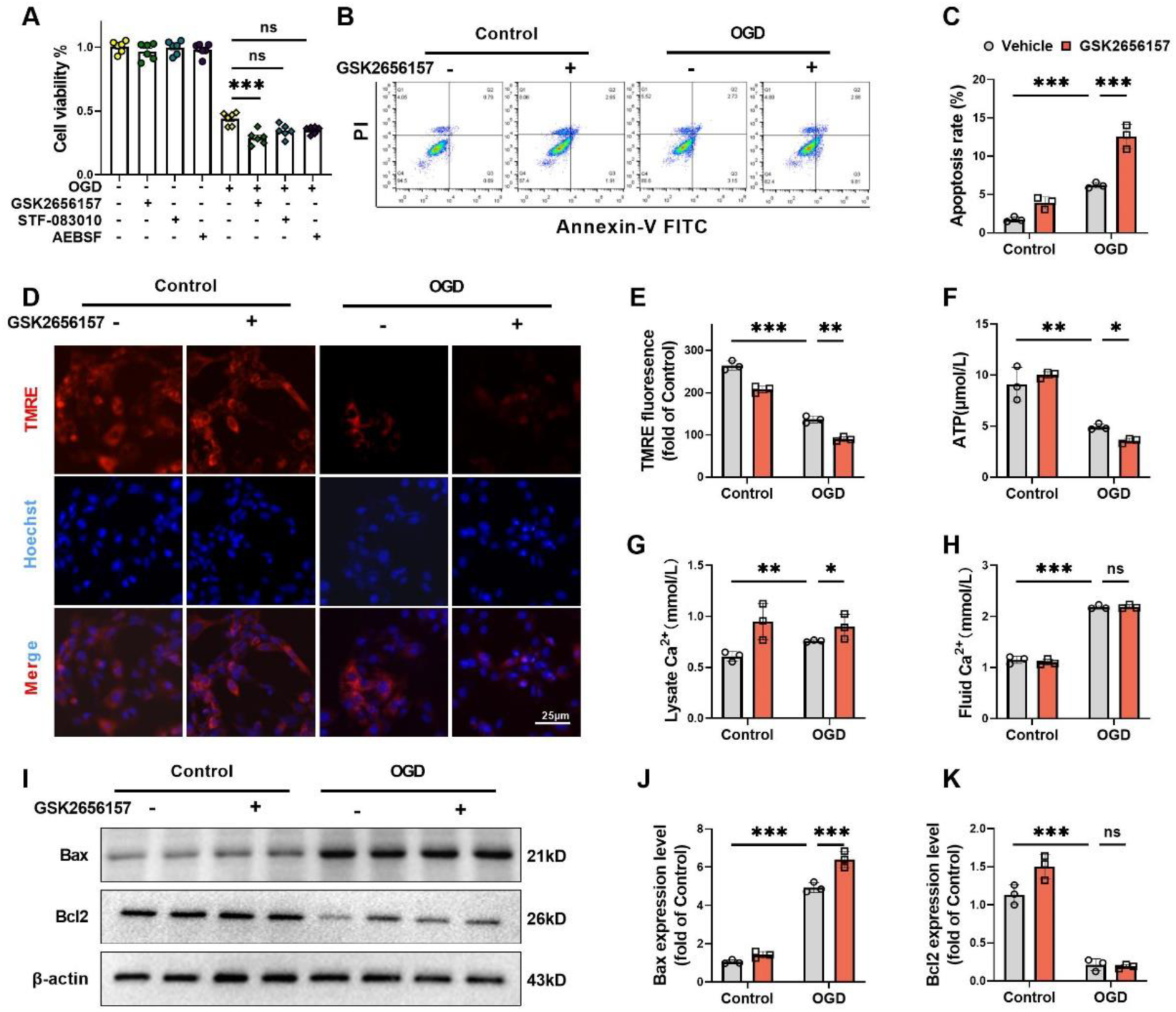
PERK has protective effects on OGD cardiomyocytes. Use GSK2656157 (10 μM), STF-083010 (25 μM), AEBSF (200 μM) inhibits endoplasmic reticulum stress and CCK8 detects cell viability in rat myocardium H9c2 cells (A). Values are the mean ± SEM. n = 6 per group. \*\*\**P* < 0.001, One way-ANOVA. H9c2 cells under 4-hours oxygen-glucose deprivation (OGD) stimulation. Apoptosis was evaluated by flow cytometry using Annexin–V fluorescein isothiocyanate (FITC)/propidium iodide (PI) staining(B-C). Fluorescence detection TMRE (D-E). A kit detected ATP content (F). A kit detected the calcium ion content in cell lysate (G) and cell supernatant (H). Immunoblotting detected apoptosis protein Bax and Bcl2 levels in H9c2 cells (I-K). Values are the mean ± SEM. n = 3 per group. \**P* < 0.05, \*\**P* < 0.01, \*\*\**P* < 0.001, Two way-ANOVA.

### ER stress activation elevates IP3R/GRP75/VDAC1 expression and calcium regulation

Tunicamycin (TM) causes accumulation of unfolded proteins in the ER and induces ER stress. Using tracers of ER and mitochondria to observe the Ca^2+^ content in two organelles after ER stress activation, we found that Ca^2+^ accumulated in the ER and increased in the mitochondria (**Figure 4A**). To study ER stress and Ca^2+^ transport, MS detected ERs and calcium-regulated proteins, including Bip and GRP75 (**Figure 4B**). Maintaining the stability of the conformation of IP3Rs after activation is essential for maintaining Ca^2+^ transport in mitochondria and cytoplasm^23^. Bip is a molecular chaperone that exists on the ER, where it stabilizes the conformation of IP3Rs^24^. Thus, it can ensure the delivery of Ca^2+^ from the ER to the mitochondria. Confocal fluorescence microscopy observed that GRP75 co-localized with IP3R and VDAC1. This colocalization was enhanced in the case of endoplasmic reticulum stress induced by TM compared to controls (**Figure 4C**). Immediately, we examined the protein expression of IP3R/GRP75/VDAC1, and the results showed that the protein expression level was increased in the TM group in likewise (**Fig. 4D-G**). Mitochondrial calcium uniporter (MCU), the calcium-binding protein on the IMM (**Fig. 4H**), was activated. And ATPase sarcoplasmic/endoplasmic reticulum Ca^2+^ transporting 2 (SERCA2) is a calcium recycling protein on the ER. After ER stress activation, both ER and calcium channel proteins on the mitochondria were activated, showing the role of maintaining calcium homeostasis.

**Figure 4.**
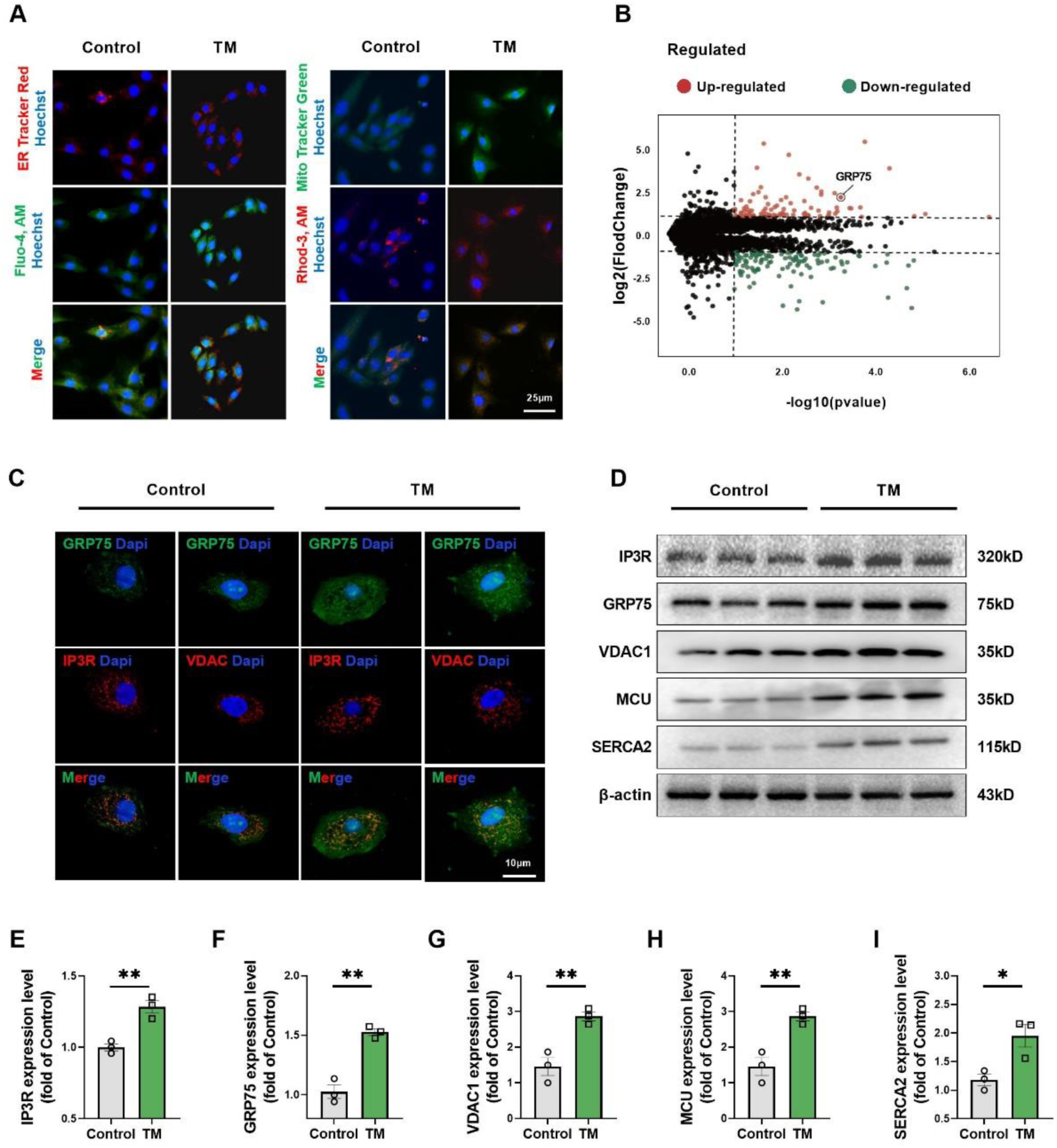
Activation of ER stress leads to higher expression of IP3R / GRP75 / VDAC1. Endoplasmic reticulum stress is activated with Tunicamycin (TM). ER Tracker Red and Mito Tracker Green marked the organelles, Fluo-4, AM and Rhod-3,AM symbolized the position of calcium ions in different organelles (A). Collecting TM-treated 24-hours H9c2 cells for mass spectrometry analysis showed a high expression of GRP75 (B). The protein content of IP3R/GRP75/VDAC1 was observed with confocal microscope (C). Immunoblotting detected protein IP3R, GRP75, VDAC1 and Ca^2+^-regulated proteins MCU and SERCA2 levels in H9c2 cells (D -I). Values are the mean ± SEM. n = 3 per group. \**P* < 0.05, \*\**P* < 0.01, two-tailed Student’s t-test.

### Inhibition of GRP75 expression rescues damaged myocardium

Szabadkai et al.^25^ suggested that GRP75 has a central role in building protein complexes with IP3R and VDAC1, suggesting that GRP75 assumes a scaffolding function in the complexes. This led us to hypothesize that GRP75 would play an important role in calcium exchange between the ER and mitochondria. We knocked down GRP75 using siRNA, and a synchronized reduction in protein expression was seen in IP3R/GRP75/VDAC1/MCU after knocking down GRP75 compared with the OGD group (**Figure 5A**). This suggests that this channel is a whole and undergoes synchronized changes when affected. Interestingly, knockdown of GRP75 did not affect the expression of hypoxia-inducible factor 1α (HIF-1α) in H9c2 cells in the face of OGD. In contrast, PERK content did not show significant changes due to knockdown of GRP75 (**Figure 5B**). This suggests that activation of IP3R/GRP75/VDAC1 occurs downstream of ER stress. Examining Ca^2+^ in the ER by Fluo-4, AM, we found that the calcium fluorescence intensity was significantly higher in the OGD group than in the Control group. And after down-regulation of GRP75 expression, ER Ca^2+^ appeared to be mildly elevated (**Figure 5C-D**). siRNA down-regulation of GRP75 expression resulted in a significant decrease in the magnitude of OGD-stimulated increase in mitochondrial Ca^2+^ relative to the OGD group (**Figure 5E**). To clarify the flow of Ca^2+^, we examined calcium in cell homogenates, and there was no doubt that OGD stimulation triggered an overall elevation of Ca^2+^ in the cells, and that down-regulation of GRP75 decreased the intracellular concentration of Ca^2+^ (**Figure 5F**). Taken together, these data suggest that inhibition of the IP3R/GRP75/VDAC1 complex prevents the transfer of calcium from the ER to mitochondria during ERS and attenuates mitochondrial calcium overload. To determine the effects of ER stress on the heart, we observed the ultrastructure of H9c2 cells by transmission electron microscopy. Neatly arranged and uniformly sized ER and mitochondria were observed in the negative control (NC) group. OGD-induced ER stress resulted in ultrastructural alterations in cardiomyocytes as evidenced by dilation of the ER, breakage of the mitochondrial cristae, and the appearance of vacuoles. In the si-GRP75-treated OGD group, only partial expansion of the endoplasmic reticulum and slight vacuolisation of the mitochondria were observed (**Fig. 5G**). OGD increased the number of contact points between the ER and the mitochondria, which reflected the formation of MAM. In the si-GRP75-treated OGD group, the number of contact points was reduced compared with the OGD group (**Fig. 5H**). In order to verify the protective effect of knockdown of GRP75 on cardiomyocytes, we used TUNEL staining to label the apoptotic cells, and the results showed that the TUNEL positivity rate of the cells in the OGD group was significantly higher compared with that of the control group, whereas si-GRP75 presented a decrease in apoptotic cells in the face of OGD (**Figure 5I-J**). Cardiomyocytes, as energy-consuming cells, require large amounts of ATP to maintain normal function. In response to OGD treatment, ATP production was substantially decreased compared with the control group. This injury was reversed by si-GRP75 (**Figure 5K**). The above results indicated that knockdown of GRP75 could regulate the formation of MAM to rescue the damaged cardiomyocytes.

**Fig 5.**
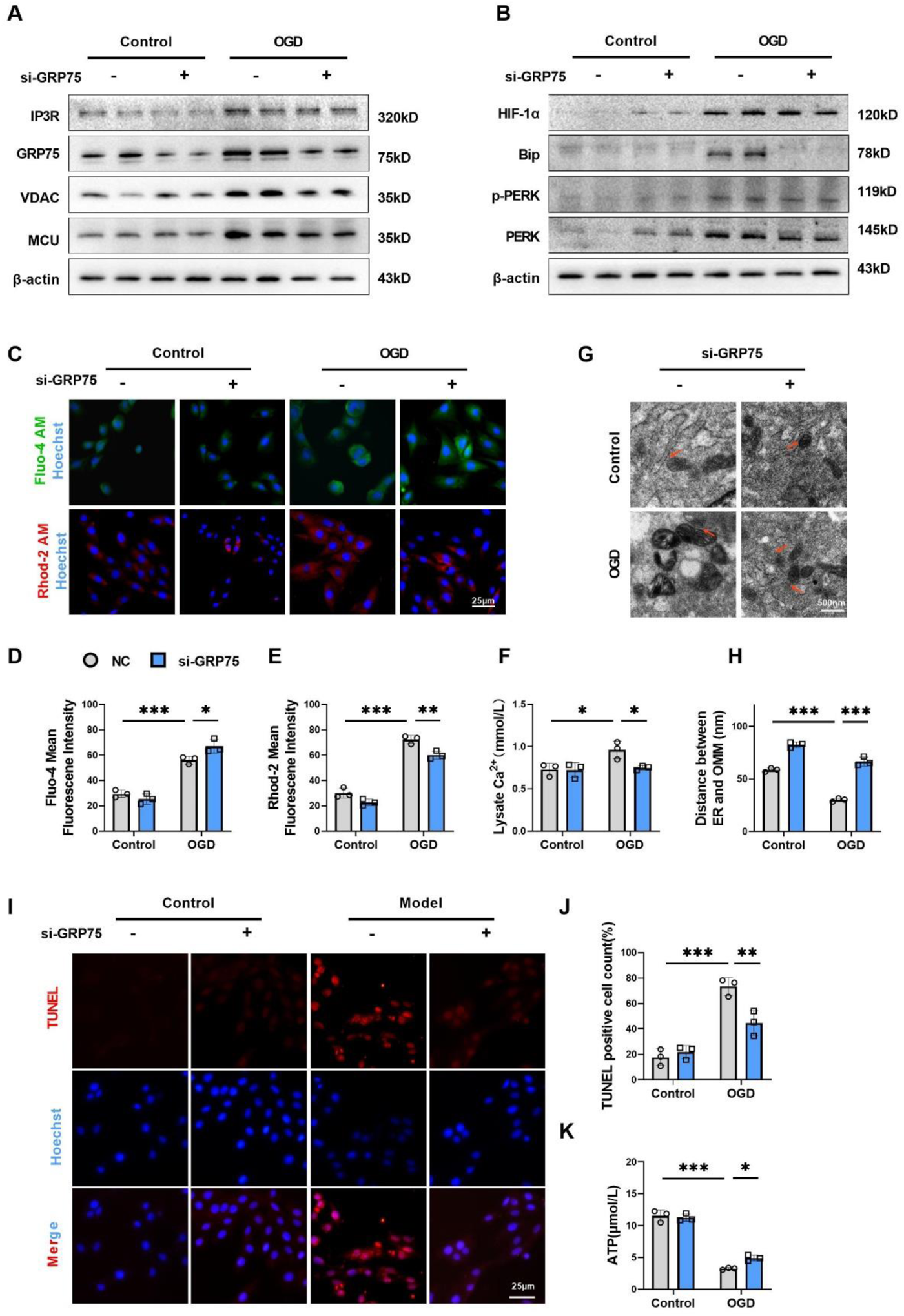
Inhibits the expression of GRP75 can rescue damaged cardiomyocytes Immunoblotting detected protein IP3R, GRP75, VDAC1 and MCU levels in H9c2 cells (A). Immunoblotting detected protein HIF-1α, Bip, p-PERK and PERK levels in H9c2 cells (B). Fluo-4, AM and Rhod-2, AM were used to detect the Ca^2+^ content (C-E) in the endoplasmic reticulum and mitochondria. A kit detected the calcium ion content in cell lysate (F). The structure of MAM was observed under a transmission electron microscope and indicated with a red arrow (G). And measured the distance between the endoplasmic reticulum and the outer mitochondrial membrane (H). Fluorescence detection of apoptosis (I-J). A kit detected ATP content in cell lysate (K). Values are the mean ± SEM. n = 3 per group. \**P* < 0.05, \*\**P* < 0.01, \*\*\**P* < 0.001, Two way-ANOVA.

### GRP75 knockdown modulates glycolysis and protects hypoxic cardiomyocytes

To elucidate the pathways through which GRP75 acts, we collected NC and siRNA knockdown GRP75 cells for transcriptome sequencing. Between-group analysis was performed using |Log2 FlodChange|≥1 and pvalue <0.05 as the screening criteria for differentially expressed genes. A total of 38 differentially expressed genes were identified, including 23 up-regulated genes and 15 down-regulated genes. Heatmap followed by volcano map was plotted according to this (**Figure 6A-C**). To further explore the effect of GRP75 knockdown on the transcriptome of H9c2 cells, the differentially expressed genes were categorized by GO Enrichment analysis, including biological process and molecular function. Biological process was mainly involved in sialylation and glycoprotein metabolism, while molecular function focused on sialyltransferase activity and IP3R activity (**Figure 6D**). Differential genes were mainly enriched in the glycospjingolipid biosynthesis-neolactate series, which can be saw in KEGG Enrichiment. (**Fig. 6E**). qPCR to detect genes of the glycolytic pathway was significantly elevated in the Model group compared with the Control group, and this elevation was eliminated after knockdown of GRP75 (**Fig. 6F**). Immediately following this, we examined the expression of related proteins and could find that glycolysis-related proteins were activated in the face of oxygen & glucose deprivation, and knocking down GRP75 attenuated this aberrant activation (**Figure 6G-K**). We then examined the levels of lactate and pyruvate, key products of glycolysis. The value of lactate/pyruvate is often used clinically to determine the degree of circulatory failure. During opposite oxygen & glucose deprivation, cardiomyocyte glycolysis was enhanced in the Model group, producing large amounts of lactate and pyruvate. Knockdown of GRP75 significantly reduced lactate/pyruvate production. Overall, inhibition of GRP75 exerts its role in rescuing hypoxic cardiomyocytes through reducing glycolysis.

**Figure 6.**
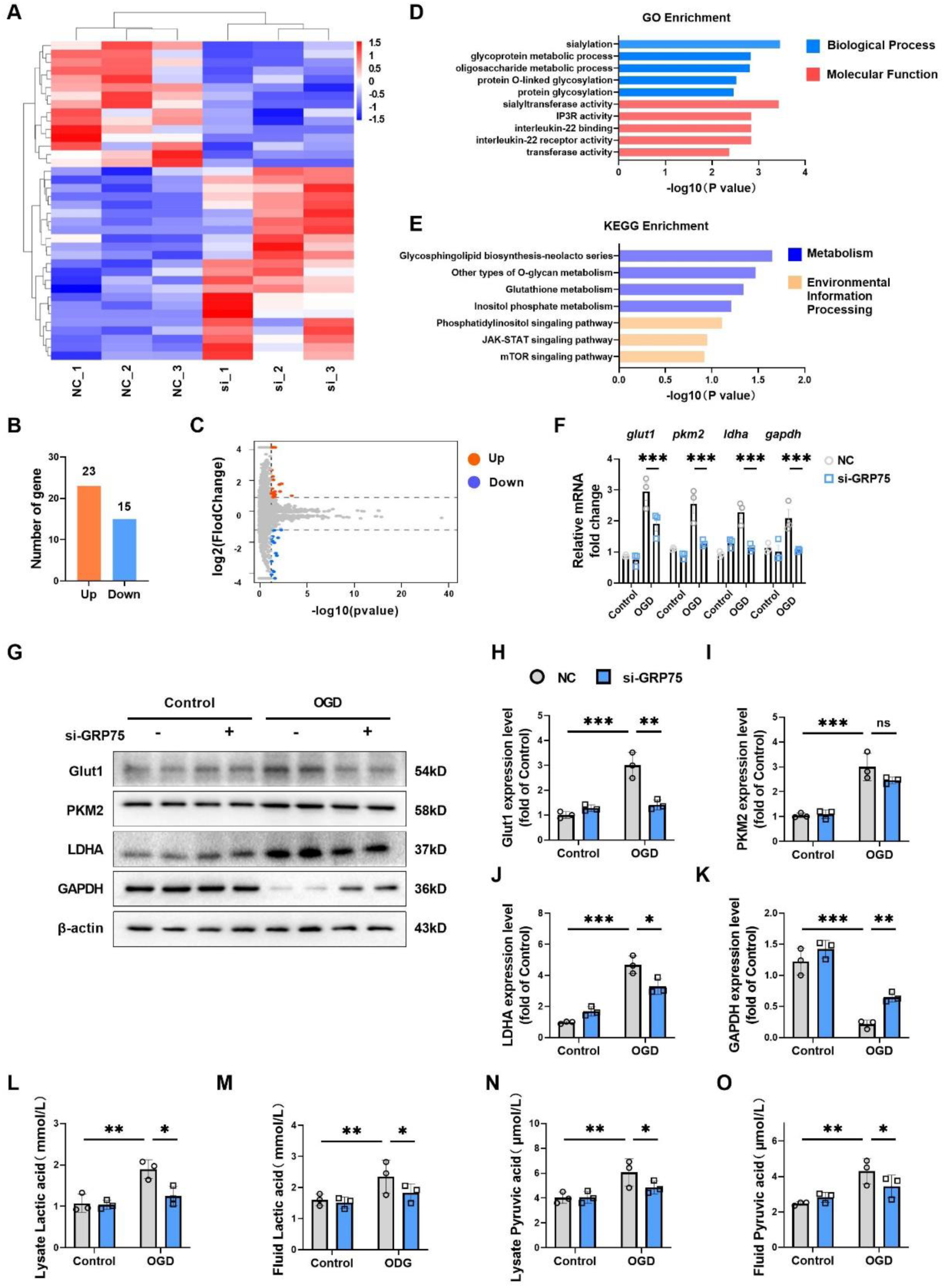
GRP75 acts through glycolysis GRP75 was silenced by siRNA in H9c2 cells, and cell samples were collected for transcriptome sequencing and compared with negative controls (NC). Transcriptome sequencing heatmap (A) with GO (B) and KEGG (C) enrichment. Differential gene number (D) with volcano plot (E). qPCR detected key enzyme activities in glycolysis gene glut1, pkm2, ldha and gapdh levels in H9c2 cells (F). Immunoblotting detected relative protein Glut1, PKM2, LDHA and GAPDH levels in H9c2 cells (G-K). A kit detects the actate and pyruvate in H9c2 cell lysate and supernatant (L-O). Values are the mean ± SEM. n = 3 per group. \**P* < 0.05, \*\**P* < 0.01, \*\*\**P* < 0.001, Two way-ANOVA.

## Discussion

Cardiovascular disease places a serious burden on the global health system. When the ER stress occurs, it may lead to myocardial infarction. The connection between mitochondria and the ER, particularly the MAM, regulates calcium homeostasis in cells and affects myocardial function. The current study proposes that GRP75 a key gene of MAM, protects mitochondrial calcium homeostasis by affecting key proteins of MAM reducing cardiomyocyte glycolysis and myocyte ischemia-hypoxia injury. More deeply, we confirmed that the IP3R1-GRP75-VDAC1 complex mediates ER stress and mitochondrial calcium disturbances after MI, and the PERK inhibitor GSK exacerbates these changes. Moreover, the knocking down of GRP75 inhibits Ca^2+^ transfer between the ER and the mitochondria, protects mitochondrial function, and attenuates cardiomyocyte injury.

The ER is an important site for regulating protein synthesis, modification, and transport in eukaryotic cells. When unfolded or misfolded proteins accumulate excessively in the ER can cause ER stress and trigger the unfolded protein response(UPR) when triggered^26^. The UPR transmits information about the folding status of proteins in the lumen of the ER to the nucleus and cytoplasm of the cell to reduce the misfolded proteins and unfolded proteins in the ER, thereby maintaining ER homeostasis^27^. Short-term UPR will to a certain degree alleviate and mitigate the burden of and damage to the ER, restore ER protein homeostasis, and long-term UPR will ensure that the cell is headed for self-destruction, which in turn leads to diseases^28^. Three typical UPR signaling pathways have been identified: the PERK, IRE1α and ATF6, which normally remain inactive by interacting with the chaperone molecule Bip (Binding immunoglobulin protein)^29^.

New evidence suggests that the three UPR signaling branches are not activated at the same time; activation of ATF6 and IRE1α occurs immediately and, through unidentified mechanisms attenuates over time^30^. Activation of PERK immediately follows activation of ATF6α and IRE1α; prolonged activation of the UPR induces apoptosis primarily through activation of the PERK pathway^31^. Minamino et al^32^ have proposed that prolonging the adaptive period of the UPR to enhance cell survival and recovery or to inhibit ER stress-associated apoptosis may help combat a range of human diseases. Previous studies have indicated that cardiac-specific PERK gene deletion exacerbates congestive heart failure caused by pressure overload^33^. In contrast, Molkentin et al^34^ found that constitutive overexpression and activation of PERK in the heart promote cardiac atrophy and lethality. Our experimental results demonstrate that Bip is linked to the activation of 3 UPR signaling pathways (PERK, ATF6, IRE1) after MI, and the activation of the PERK pathway was particularly prominent. In addition, we found that inhibition of the PERK pathway causes more severe cardiomyocyte damage.

MAMs are membrane contact sites between mitochondria and the ER that bi-directionally regulate organelle physiological functions such as calcium and lipid homeostasis, mitochondrial dynamics, and apoptosis^35^. The key protein for calcium ion efflux from the ER is the IP3R, which is physically linked to the VDACs on the OMM via GRP75 to constitute a tripartite complex. The IP3R1-GRP75-VDAC1 complex forms a Ca^2+^ regulatory axis with the MCU, which mediates Ca^2+^ transport from the ER to the mitochondria^36^. Ca^2+^ is a second messenger that regulates cellular metabolism and apoptosis and other multiple cellular activities. As in the current study, we extracted MI transcriptome data from the GEO database in Chinese males and found that after MI, most of the genes were differentially expressed in relation to metabolic pathways, with the calcium signaling pathway being particularly altered.

MCU-mediated mitochondrial calcium influx plays an important role in inducing mitochondrial permeability transition pore (mPTP) opening and cell death under pathological conditions such as ischemia/reperfusion^37^. The mitochondrial calcium influx is a key component of mPTP opening and cell death. Melanie et al^38^ reported that Ca^2+^ in the ER flows to the mitochondria via the VDAC1/GRP75/IP3R1 complex during the onset of myocardial ischemia-reperfusion. This resulted in mitochondrial calcium overload, triggering mitochondria-dependent apoptosis. In contrast, calcium flow from the ER to mitochondria can be inhibited by inhibiting VDAC1 or MFN2, thereby attenuating ischemia-reperfusion injury in the myocardium. We found that GRP75 plays a central role in forming a protein complex with IP3R and VDAC1 that affects calcium exchange between the ER and mitochondria. After the knockdown of GRP75, the protein expression of IP3R, GRP75, VDAC1, and MCU was synchronously reduced, suggesting that the channel functions as a tight unit to mediate cardiomyocyte injury. More importantly, PERK levels were unaffected after the knockdown of GPR75, suggesting that activation of the IP3R/GRP75/VDAC1 complex is a downstream event after endoplasmic reticulum stress and that PERK may mediate the formation of the IP3R/GRP75/VDAC1 complex.

In mouse primary neurons, GRP75 promotes ER-mitochondrial association and increases ATP production for axonal regeneration^39^. In type 2 diabetic mice, the knockdown of GRP75 ameliorates ERs-induced atrial remodeling and reduces the development of atrial fibrillation. We found that when ER stress occurs in cardiomyocytes, the expression of IP3R, GRP75, and VDAC1 all rise, and that these proteins play a key role in the maintenance of intracellular calcium homeostasis. By examining calcium levels, we found that inhibition of the IP3R/GRP75/VDAC1 complex blocked calcium transfer from the ER to mitochondria during ER stress and subsequently reduced mitochondrial calcium overload. We suggest that ER and mitochondria in cardiomyocytes are in close proximity to each other in the face of OGD, followed by translocation of calcium ions from the endoplasmic reticulum lumen and across the OMM through the MCU into mitochondria via the IP3R/GRP75/VDAC1 complex.

The adult mammalian heart utilizes fatty acids as its primary energy source, and in heart failure fatty acid metabolism decreases and glucose metabolism increases^40^. Glucose is taken up by glucose transporter proteins and enters the cardiomyocyte, where it undergoes glycolysis to produce pyruvate and lactate. Most of the pyruvate enters the mitochondria and is converted to acetyl coenzyme A, which enters the TCA cycle and is metabolized to produce ATP. ATP produced by anaerobic glycolysis of glucose accounts for 2% to 8% of the ATP produced by the mitochondria of cardiomyocytes^41^. Our sequencing revealed that the knockdown of GRP75 in normal cardiomyocytes resulted in changes in the expression of glycolysis-related genes and proteins and significantly reduced the production of lactate and pyruvate, both of which are key products of glycolysis. The results of our experiments enriched the differences brought about by knocking down the GRP75 gene in the sugar metabolism pathway. Immediately thereafter, we verified that knocking down GRP75 improves glycolysis in OGD cardiomyocytes and increases ATP production to improve cardiac function. These suggest that the GRP75 inhibiting can rescue cardiomyocyte apoptosis induced by ER stress, potentially by reducing glycolysis.

In summary, the intricate interplay between the ER and mitochondria, particularly mediated by GRP75 and the associated IP3R1-GRP75-VDAC1 complex, plays a pivotal role in calcium homeostasis and the cardiomyocyte response to ER stress. The current study underscores the significance of GRP75 in the regulation of intracellular calcium dynamics and its influence on glycolytic pathways in cardiomyocytes, providing a novel perspective on the possible therapeutic targets for mitigating myocardial injuries by reducing ER stress. While our results are promising and shed light on potential therapeutic avenues, the translatability of these findings to in vivo conditions, especially in diverse human populations, remains to be validated. Additionally, while we focused on GRP75’s role in calcium homeostasis and glycolysis, it’s plausible that other molecules or pathways may be co-contributors. Future investigations would benefit from a more comprehensive in vivo study design, and a wider exploration of other potential molecules involved, ensuring a holistic understanding of the intricate cellular response post-myocardial infarction.

## Conclusions

Following myocardial infarction ER stress is activated and MAM mediates Ca^2+^ delivery from the endoplasmic reticulum to mitochondria. The key proteins on MAM, IP3R/GRP75/MCU, provide a dependent channel for Ca^2+^ delivery. Knockdown of GRP75 expression reduces calcium ion accumulation in mitochondria, cardiomyocyte injury, and acts by regulating glycolysis.

## Data Availability

The datasets used and/or analyzed during the current study are available from the corresponding author on reasonable request.

## Sources of Funding

This study was supported by the National Natural Science Foundation of China (No. 82204382; 81500310), supporting project of National Natural Science Foundation of Nanjing University of Chinese Medicine (XPT82204382), Jiangsu Key Discipline Construction Fund of the 14th Five-Year Plan (Biology).

## Contributions

CYZ took part in all the experiments. BWL and JXS composed the manuscript. WJZ and JW raised the rats. CX, XZ and YXZ gathered all the data. QHM and YL contributed to the design of the study and the writing of the manuscript. QHM and YL received support from the funds and supervised the study.

## Ethics declarations

The animal experiments were approved by the Experimental Animal Ethics Committee of Nanjing University of Chinese Medicine (Animal ethic NO. 201912A012).

## Consent for publication

Not applicable for that section.

## Competing interests

The authors declare that they have no competing interests.

---

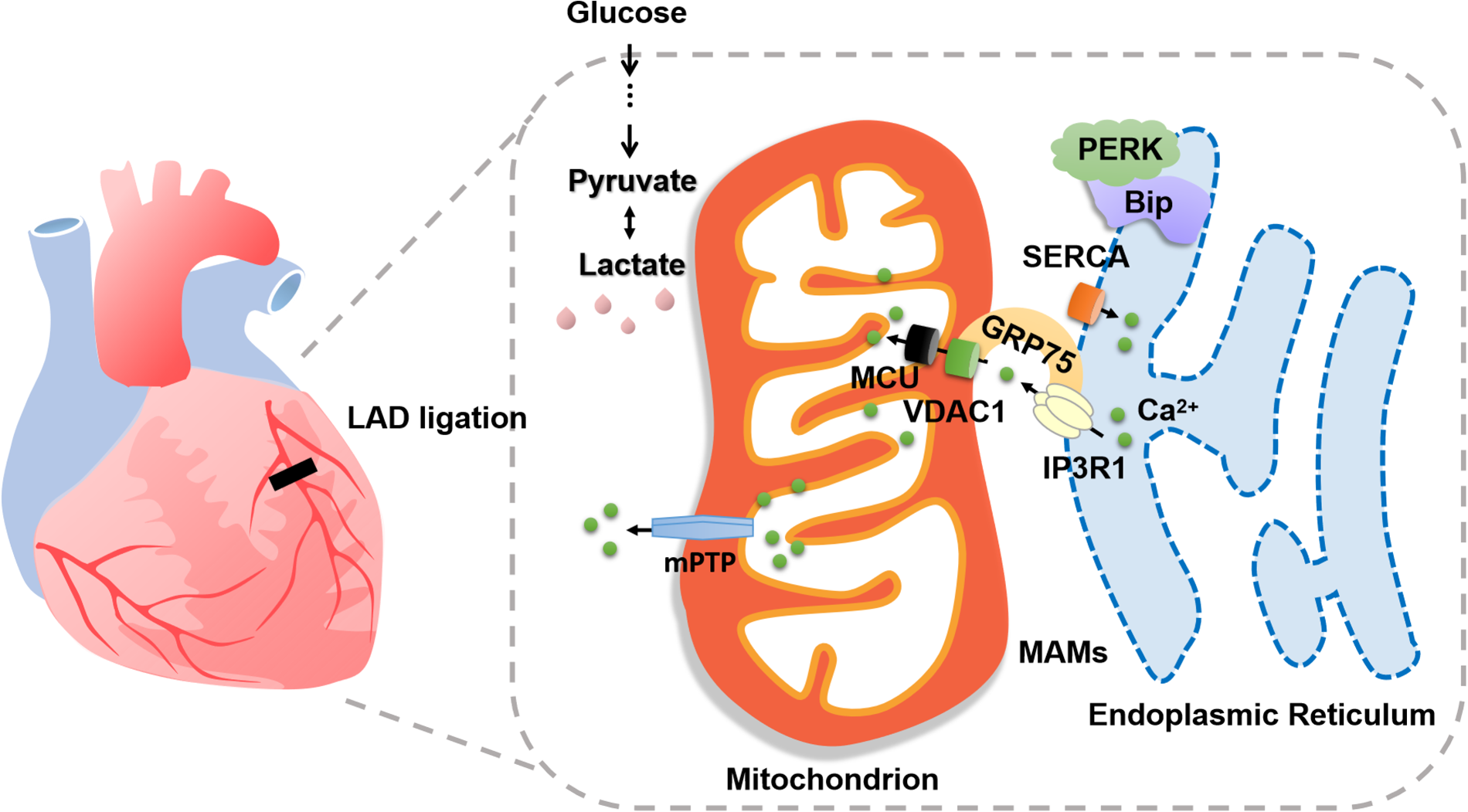

## Notes

### Competing Interest Statement

The authors have declared no competing interest.

### Author Declarations

Nanjing University of Chinese Medicine

## References

1. Stretti L, Zippo D, Coats AJS, Anker MS, von Haehling S, Metra M, Tomasoni D. A year in heart failure: an update of recent findings. ESC Heart Fail. 2021;8:4370–4393.

2. Wang M, Kaufman RJ. Protein misfolding in the endoplasmic reticulum as a conduit to human disease. Nature. 2016;529:326–335.

3. Jiang Y, Tao Z, Chen H, Xia S. Endoplasmic Reticulum Quality Control in Immune Cells. Front Cell Dev Biol. 2021;9:740653.

4. Patergnani S, Suski JM, Agnoletto C, Bononi A, Bonora M, De Marchi E, Giorgi C, Marchi S, Missiroli S, Poletti F, Rimessi A, Duszynski J, Wieckowski MR, Pinton P. Calcium signaling around Mitochondria Associated Membranes (MAMs). Cell Commun Signal CCS. 2011;9:19.

5. Schwarz DS, Blower MD. The endoplasmic reticulum: structure, function and response to cellular signaling. Cell Mol Life Sci. 2016;73:79–94.

6. Paillard M, Tubbs E, Thiebaut P-A, Gomez L, Fauconnier J, Da Silva CC, Teixeira G, Mewton N, Belaidi E, Durand A, Abrial M, Lacampagne A, Rieusset J, Ovize M. Depressing mitochondria-reticulum interactions protects cardiomyocytes from lethal hypoxia-reoxygenation injury. Circulation. 2013;128:1555–1565.

7. De Nicolo B, Cataldi-Stagetti E, Diquigiovanni C, Bonora E. Calcium and Reactive Oxygen Species Signaling Interplays in Cardiac Physiology and Pathologies. Antioxid Basel Switz. 2023;12:353.

8. Pedriali G, Ramaccini D, Bouhamida E, Wieckowski MR, Giorgi C, Tremoli E, Pinton P. Perspectives on mitochondrial relevance in cardiac ischemia/reperfusion injury. Front Cell Dev Biol. 2022;10:1082095.

9. Giorgi C, Marchi S, Pinton P. The machineries, regulation and cellular functions of mitochondrial calcium. Nat Rev Mol Cell Biol. 2018;19:713–730.

10. Perrone M, Caroccia N, Genovese I, Missiroli S, Modesti L, Pedriali G, Vezzani B, Vitto VAM, Antenori M, Lebiedzinska-Arciszewska M, Wieckowski MR, Giorgi C, Pinton P. The role of mitochondria-associated membranes in cellular homeostasis and diseases. Int Rev Cell Mol Biol. 2020;350:119–196.

11. Janikiewicz J, Szymański J, Malinska D, Patalas-Krawczyk P, Michalska B, Duszyński J, Giorgi C, Bonora M, Dobrzyn A, Wieckowski MR. Mitochondria-associated membranes in aging and senescence: structure, function, and dynamics. Cell Death Dis. 2018;9:332.

12. Li YE, Sowers JR, Hetz C, Ren J. Cell death regulation by MAMs: from molecular mechanisms to therapeutic implications in cardiovascular diseases. Cell Death Dis. 2022;13:504.

13. Xu H-X, Cui S-M, Zhang Y-M, Ren J. Mitochondrial Ca^2+^ regulation in the etiology of heart failure: physiological and pathophysiological implications. Acta Pharmacol Sin. 2020;41:1301–1309.

14. Woll KA, Van Petegem F. Calcium-release channels: structure and function of IP3 receptors and ryanodine receptors. Physiol Rev. 2022;102:209–268.

15. Szabadkai G, Bianchi K, Várnai P, De Stefani D, Wieckowski MR, Cavagna D, Nagy AI, Balla T, Rizzuto R. Chaperone-mediated coupling of endoplasmic reticulum and mitochondrial Ca2+ channels. J Cell Biol. 2006;175:901–911.

16. Yuan M, Gong M, He J, Xie B, Zhang Z, Meng L, Tse G, Zhao Y, Bao Q, Zhang Y, Yuan M, Liu X, Luo C, Wang F, Li G, Liu T. IP3R1/GRP75/VDAC1 complex mediates endoplasmic reticulum stress-mitochondrial oxidative stress in diabetic atrial remodeling. Redox Biol. 2022;52:102289.

17. Li Y, Li H-Y, Shao J, Zhu L, Xie T-H, Cai J, Wang W, Cai M-X, Wang Z-L, Yao Y, Wei T-T. GRP75 Modulates Endoplasmic Reticulum-Mitochondria Coupling and Accelerates Ca2+-Dependent Endothelial Cell Apoptosis in Diabetic Retinopathy. Biomolecules. 2022;12:1778.

18. Xu H, Guan N, Ren Y-L, Wei Q-J, Tao Y-H, Yang G-S, Liu X-Y, Bu D-F, Zhang Y, Zhu S-N. IP3R-Grp75-VDAC1-MCU calcium regulation axis antagonists protect podocytes from apoptosis and decrease proteinuria in an Adriamycin nephropathy rat model. BMC Nephrol. 2018;19:140.

19. Wang T, Zhu Q, Cao B, Cai Y, Wen S, Bian J, Zou H, Song R, Gu J, Liu X, Liu Z, Yuan Y. Ca2+ transfer via the ER-mitochondria tethering complex in neuronal cells contribute to cadmium-induced autophagy. Cell Biol Toxicol. 2022;38:469–485.

20. Liu X-H, Zhang Z-Y, Sun S, Wu X-D. Ischemic postconditioning protects myocardium from ischemia/reperfusion injury through attenuating endoplasmic reticulum stress. Shock Augusta Ga. 2008;30:422–427.

21. Li B, Tian J, Sun Y, Xu T-R, Chi R-F, Zhang X-L, Hu X-L, Zhang Y-A, Qin F-Z, Zhang W-F. Activation of NADPH oxidase mediates increased endoplasmic reticulum stress and left ventricular remodeling after myocardial infarction in rabbits. Biochim Biophys Acta. 2015;1852:805–815.

22. Martindale JJ, Fernandez R, Thuerauf D, Whittaker R, Gude N, Sussman MA, Glembotski CC. Endoplasmic reticulum stress gene induction and protection from ischemia/reperfusion injury in the hearts of transgenic mice with a tamoxifen-regulated form of ATF6. Circ Res. 2006;98:1186–1193.

23. Hayashi T, Su T-P. Sigma-1 receptor chaperones at the ER-mitochondrion interface regulate Ca(2+) signaling and cell survival. Cell. 2007;131:596–610.

24. Higo T, Hamada K, Hisatsune C, Nukina N, Hashikawa T, Hattori M, Nakamura T, Mikoshiba K. Mechanism of ER stress-induced brain damage by IP(3) receptor. Neuron. 2010;68:865–878.

25. Szabadkai G, Bianchi K, Várnai P, De Stefani D, Wieckowski MR, Cavagna D, Nagy AI, Balla T, Rizzuto R. Chaperone-mediated coupling of endoplasmic reticulum and mitochondrial Ca2+ channels. J Cell Biol. 2006;175:901–911.

26. Tang Q, Liu Q, Li Y, Mo L, He J. CRELD2, endoplasmic reticulum stress, and human diseases. Front Endocrinol. 2023;14:1117414.

27. Senft D, Ronai ZA. UPR, autophagy, and mitochondria crosstalk underlies the ER stress response. Trends Biochem Sci. 2015;40:141–148.

28. Herrema H, Guan D, Choi JW, Feng X, Salazar Hernandez MA, Faruk F, Auen T, Boudett E, Tao R, Chun H, Ozcan U. FKBP11 rewires UPR signaling to promote glucose homeostasis in type 2 diabetes and obesity. Cell Metab. 2022;34:1004–1022.e8.

29. Wang M, Kaufman RJ. Protein misfolding in the endoplasmic reticulum as a conduit to human disease. Nature. 2016;529:326–335.

30. Lin JH, Li H, Yasumura D, Cohen HR, Zhang C, Panning B, Shokat KM, Lavail MM, Walter P. IRE1 signaling affects cell fate during the unfolded protein response. Science. 2007;318:944–949.

31. Rutkowski DT, Arnold SM, Miller CN, Wu J, Li J, Gunnison KM, Mori K, Sadighi Akha AA, Raden D, Kaufman RJ. Adaptation to ER stress is mediated by differential stabilities of pro-survival and pro-apoptotic mRNAs and proteins. PLoS Biol. 2006;4:e374.

32. Minamino T, Komuro I, Kitakaze M. Endoplasmic reticulum stress as a therapeutic target in cardiovascular disease. Circ Res. 2010;107:1071–1082.

33. Liu X, Kwak D, Lu Z, Xu X, Fassett J, Wang H, Wei Y, Cavener DR, Hu X, Hall J, Bache RJ, Chen Y. Endoplasmic reticulum stress sensor protein kinase R-like endoplasmic reticulum kinase (PERK) protects against pressure overload-induced heart failure and lung remodeling. Hypertens Dallas Tex 1979. 2014;64:738–744.

34. Vanhoutte D, Schips TG, Vo A, Grimes KM, Baldwin TA, Brody MJ, Accornero F, Sargent MA, Molkentin JD. Thbs1 induces lethal cardiac atrophy through PERK-ATF4 regulated autophagy. Nat Commun. 2021;12:3928.

35. Zhang Y, Yao J, Zhang M, Wang Y, Shi X. Mitochondria-associated endoplasmic reticulum membranes (MAMs): Possible therapeutic targets in heart failure. Front Cardiovasc Med. 2023;10:1083935.

36. Xu H, Guan N, Ren Y-L, Wei Q-J, Tao Y-H, Yang G-S, Liu X-Y, Bu D-F, Zhang Y, Zhu S-N. IP3R-Grp75-VDAC1-MCU calcium regulation axis antagonists protect podocytes from apoptosis and decrease proteinuria in an Adriamycin nephropathy rat model. BMC Nephrol. 2018;19:140.

37. Bertero E, Maack C. Calcium Signaling and Reactive Oxygen Species in Mitochondria. Circ Res. 2018;122:1460–1478.

38. Paillard M, Tubbs E, Thiebaut P-A, Gomez L, Fauconnier J, Da Silva CC, Teixeira G, Mewton N, Belaidi E, Durand A, Abrial M, Lacampagne A, Rieusset J, Ovize M. Depressing mitochondria-reticulum interactions protects cardiomyocytes from lethal hypoxia-reoxygenation injury. Circulation. 2013;128:1555–1565.

39. Lee S, Wang W, Hwang J, Namgung U, Min K-T. Increased ER-mitochondria tethering promotes axon regeneration. Proc Natl Acad Sci U S A. 2019;116:16074–16079.

40. Gupta A, Houston B. A comprehensive review of the bioenergetics of fatty acid and glucose metabolism in the healthy and failing heart in nondiabetic condition. Heart Fail Rev. 2017;22:825–842.

41. Lopaschuk GD, Karwi QG, Tian R, Wende AR, Abel ED. Cardiac Energy Metabolism in Heart Failure. Circ Res. 2021;128:1487–1513.

